# COVID-19 exposure risks and protective measures in East London’s healthcare and academic sectors: Insights and applications of GloBody technology for infectious disease monitoring

**DOI:** 10.1101/2025.04.17.25325996

**Authors:** Sharmilee Gnanapavan, Mohammad Aboulwafa, Francesca Ammoscato, Michael Andrews, Georgios Alampritis, A. Nazli Asardag, David Baker, Randy Chance, Carrie Chew, Teresa Cutino-Moguel, Anastasia Georgievskaya, Upkar S. Gill, Gavin Giovannoni, Charalambos Hadjicharalambous, Anna He, David W. Holden, Meleri Jones, Madeleine R. Jones, Patrick T. F. Kennedy, Ewan Main, Olivia R. McIver, Triska Miran, Adrienne Morgan, Ankit Patel, Ruth S. Rose, Klaus Schmierer, Justyna Skonieczna, Angray S. Kang, Paul L. Ryan

**Author notes:** **Correspondence**: Dr. Sharmilee Gnanapavan. Neurology, The Royal London Hospital, Barts Health NHS Trust, Mile End Road, Whitechapel, London E1 1FR, United Kingdom. Joint senior authors (Sharmilee Gnanapavan, Angray S. Kang, Paul L. Ryan).

## Abstract

The COVID-19 pandemic highlighted the need for effective protection and rapid development of tests to track and quantify seroconversion through natural infection and vaccination. Recombinant proteins, consisting of the SARS-CoV-2 nucleocapsid and Spike Receptor Binding Domains (RBD) fused with nanoluciferase reporters (GloBodies) were designed and produced. The SARS-CoV-2 specific antibody within serum, from venous blood or eluted from local or remotely-obtained dried blood spots, form a complex with the GloBody, which can be captured on immobilized Protein G or anti-isotype antibody with the retained nanoluciferase activity being proportional to specific antibody levels. Natural infection, vaccination and human and animal SARS-CoV-2 specific antibodies were detectable. These were used to serially monitor infection and vaccination responses in dental healthcare workers (n=82), medical healthcare workers (n=72) and laboratory-based scientists (n=62) within the Royal London, dental and medical hospitals and associated university research institute in Whitechapel, East London. This indicated temporally distinct infection and vaccination profiles, consistent with hospital deployments and local and national lockdowns by dentists and scientists. As such, medical healthcare workers had twice the odds of experiencing COVID-19 symptoms (2.01 95%CI 1.13- 3.58. P<0.001) compared to dentists, who were more at risk than scientists. Likewise, those who performed virus exposure-prone procedures exhibited twice (1.98. 95% CI 1.18-3.34 P=0.01) the odds of symptoms and exhibited higher nucleocapsid titres (0.21 95%CI 0.05-0.38. P=0.012), indicative of higher infection levels. As predicted vaccination was associated with reduced infection risk as shown by reduced titre of nucleocapsid titres (-0.21 95% CI -0.34- - 0.17. P<0.001 and elevated Log_10_ RBD GloBody titres (1.18. 95%CI 1.09-1.26. P<0.001). The GloBody technology proved to be a versatile and scalable platform for rapid deployment.

## INTRODUCTION

The coronavirus-2019 (COVID-19) pandemic due to Severe Acute Respiratory Syndrome Coronavirus Two (SARS-CoV-2) infection caused and continues to cause significant global human mortality, notably until mass vaccination protected populations [Zhu et al. 2020; Zhou et al. 2020; Polatoğlu et al. 2023]. In London, the heart of the COVID-19 pandemic was East London, particularly in areas like Barking and Dagenham, Newham, and Redbridge, which reported some of the highest infection rates in the city [Mackintosh 2020; Apea et al. 2021]. By early 2021, these boroughs saw infection rates around 1,300-1,500 cases per 100,000 people, among the highest in the United Kingdom (UK) at that time [Mackintosh 2020]. The Barking, Havering and Redbridge NHS Trust, managing Queen’s and King George hospitals, recorded 1,153 deaths, while Barts NHS Trust, overseeing hospitals including Whipps Cross and Royal London, reported 1,150 deaths [Mackintosh 2020].

In 2016, Exercise Alice simulated a large-scale Middle East Respiratory syndrome (MERS) outbreak in England, caused by MERS-CoV, a similarly deadly infectious disease, to assess the UK’s response capability. The exercise highlighted the need for adequately trained professionals, ample personal protection equipment (PPE), bed capacity, and specialized equipment in the early stages of an outbreak [Hallett. 2024]. However, the urgency of these warnings went unrealised till the emergence of SARS-CoV-2. The pandemic underscored critical PPE shortages, especially during the initial outbreak, impacting frontline workers’ safety. Many healthcare workers, particularly in high-risk settings, reported insufficient PPE, reflecting the extraordinary surge in demand across England in 2020 [National Audit Office. 2020].

Early during the pandemic there was a lack of assays to monitor infection. These focused on deoxyribonucleic acid (DNA) technology and polymerase chain reaction as antibody-based infection-related, diagnostic tests take time to produce and were not available. GloBody^TM^-based reagents were originally designed to monitor anti-drug antibody (ADA) responses to protein-based therapeutics used in multiple sclerosis [Saxena et al. 2020; Baker et al. 2020]. Genetically tagging the molecule to be detected with nanoluciferase produces a labelled antigen. Antibodies that recognise the antigen form a complex which can be captured on immobilized Protein G or anti-isotype antibody. The retained nanoluciferase can be quantified with the light generated being directly proportional to the amount of specific antibody in the sample. The reference “Wuhan” SARS-CoV-2 DNA sequence was rapidly disclosed and coded only four structural proteins [Zhou et al. 2020; Polatoğlu et al. 2023]. The nucleocapsid (NUC) is the major immunogenic, intraviral protein and the viral surface displayed Spike receptor S1 subunit Binding Domain (RBD) proteins required for infection were selected as GloBody fusion targets [Zhou et al. 2020; Guo et al. 2020; Polatoğlu et al. 2023]. We designed SARS-CoV-2-specific GloBody assays to detect infection and future vaccination responses.

There has been a very large number of cross-sectional studies relating SAR-CoV-2 infection and vaccine responses of medical healthcare workers (HCW) in the UK and elsewhere [Martin et al. 2022; Galgut et al. 2024], there are relatively few studies examining the infection and impact on dentistry [Shields et al. 2021;Sabbagh et al. 2022; van der Plaat et al 2022; Schwarz et al. 2024; van Capelleveen et al. 2024; Santigli et al. 2025], or the interactions between workforce in hospital, dental, and university settings during an outbreak. This study describes the production of novel anti-viral GloBody reagents used to serially monitor differences in infection rates and vaccination responses between dental and medical HCW and laboratory-based scientists working within The Royal London Hospital, University Campus, East London during the COVID-19 pandemic, where it was anticipated that people performing exposure-prone procedures and subsequently being vaccinated would develop elevated SAR-CoV-2- specific immune responses.

## MATERIALS & METHODS

### Design

The study was a multicentre longitudinal observational cohort study during the SAR- CoV-2 alpha (Lineage B.1.1.7) and delta (Lineage B.1.617.2) waves of COVID-19 in London between 2020-2021.

### Ethical Approval

The study was conducted in accordance with applicable guidelines of Good Clinical Practice under UK Clinical Trial Regulations (SI2004, 1031, as amended). Written informed consent was obtained from each participant following Ethical Approval under Integrated Research Application System numbers IRAS 285071 and IRAS 284940 and Research Ethics Committee numbers 20/HRA/2743 and 20/NE/0176. Additional pre-vaccination, diagnostically-tested samples were obtained with ethical approval 20/HRA/2675, ISRCTN15634328 and blindly analysed [Choudhry et al. 2021]

### Settings

The Royal London Hospital and the related temporary Nightingale Hospital London were major front-line healthcare and vaccination centres during the COVID-19 pandemic for East London. This serviced a very ethnically and wealth-diverse population, notably people with a Black and Asian background with disproportionate rates of premature death and morbidity from COVID-19 [Apea et al. 2021]. Dental Workers were recruited from the Royal London Dental Hospital. Aerosol-generating activities were suspended during the national COVID-related lockdowns, notably the first national lockdown in late March 2020 and staff were redeployed to assist non-dental, medical activities in the first 8 weeks of the pandemic (March 2020). Dental aerosol generating procedures (AGPs) re-started to manage dental emergencies at the Royal London Dental Hospital in May 2020 with an approximate 50% reduction in regular AGP clinical dental activity from July 2020 until February 2022 due to challenges in the use of dental units in the open plan hospital clinical environment. Normal levels of clinical AGP dental activity resumed in February 2022. Medical HCWs were recruited from the Royal London Hospital, where many medical HCWs were seconded from their standard practice to assist the COVID-19 response. Scientists were recruited from the Blizard Institute, which is an open-plan university research laboratory attached to the hospitals. However, access, which is dependent on public transport to reach the building, was largely restricted, except for essential worker and COVID- 19 related activities, from the first national lock-down in Late-March 2020 until the two metre, UK Government rule for social distancing was lifted in late June 2020 and again limited during the second (November 2020) and third National and Local Lockdowns from Early November 2020-March 2021. The frequency of COVID cases in London was obtained from the https://ukhsa-dashboard.data.gov.uk/

### Participants

Volunteers of dental HCW (Dentists, dental hygiene therapists, dental nurses. Planned n=80) and Scientists (Students, academic, technical staff) and medical HCW (Doctors, nurses, allied healthcare therapists, healthcare assistants. Planned total n=120) within the Royal London Hospital campus, aged 20-75 years were recruited if they provided informed written consent. No sample size predictions were available. No formal exclusion criteria were made, provided individuals could give informed consent and have blood samples safely drawn.

### Demographic Outputs

A questionnaire on factors that may influence SARS-CoV-2 infection and immunity, including age, gender, ethnicity (classified as White, Black, Asian, Mixed or Other per UK census categories), COVID-19-related symptoms (see below), exposure-prone procedures and vaccination were collected at the visits. Anti-SARS-CoV-2 seropositivity and neutralizing antibody status, questionnaire findings were stored using pseudo anonymised identifiers in the National Health Service (NHS)/University computer by the Chief Investigator (SG) and immediate research staff performing the analysis, including the statistician. The source data identifying the participant was only stored on an NHS computer by the Chief Investigator.

### Exposure Variables

The main exposures of interest were 1. Occupation type, and 2. Exposure-prone procedures. Occupation type was a 3-level factor variable: dental, non-dental medical healthcare worker and scientists. Exposure-prone procedures were binary variables (yes/no) that were ascertained by participant self-report at each study visit, indicating whether exposure-prone procedures were performed between the last study visit and the current study visit. In the case of the first study visit, this indicated whether the participant performed exposure-prone procedures during the pandemic period preceding the baseline study visit. Covariates included in outcome analyses included age at baseline study visit, gender, vaccination status at time of outcome measurement. Descriptive analyses also included vaccine type received, timing of vaccinations, history of symptoms prior to study start, and race/ethnicity (categorized as White, Black, Asian, Mixed and Other according to the UK census categories) *Outcomes*. Three outcomes were included in longitudinal analyses: 1. Nucleocapsid Assay titres (relative light units) 2. RBD Assay titres (relative light units), and 3. The incidence of symptoms was ascertained at each study visit. Occurrence of COVID-19 symptoms (yes/no) was ascertained by self-report at each study visit, indicating the occurrence of at least one of the following symptoms between the previous and current visit (and not attributable to a more likely diagnosis): fever/chills, runny nose, cough, dyspnea, sore throat, headache, fatigue, muscle pain, diarrhoea, new loss of taste or smell, or other COVID-related symptom [Zhu et al. 2020]. Polymerase chain reaction confirmation of infection was typically not available. In the case of the first study visit, this indicated whether the participant experienced COVID-19-like symptoms during the pandemic period preceding the baseline study visit.

### Samples

For this study, serum was prospectively collected from 10mL of blood from dental workers who provided samples at two monthly intervals for at least 12 months. Monthly samples for 6 months were collected from medical HCW and scientists. Serum was processed in safety cabinets and the virus was inactivated by adding equal volumes of PBS 2% Triton-X-100 [Darnell & Taylor 2006; Welch et al. 2020]. Samples were stored at -20°C until used.

In addition, dried capillary blood spots were collected using lancets, Whatman 903 Protein Saver cards and used as part of assay development for remote testing [Tallantyre et al. 2022]. Blood spots were stored desiccated at room temperature and reconstituted by punching a 4mm disk into 0.2mL PBS 1% Triton-X-100. Venous serum samples from SARS-CoV-2-infected and non-infected individuals that had been tested using SARS-CoV-2 diagnostic serological assays [Choudhry et al. 2021] were also used for assay development. These were tested using the Roche Elecsys® anti-SARS-CoV-2 Pan Ig assay and the Roche Cobas e801 analyser [Choudhry et al. 2021; Gaeta et al. 2023], which is a semi-quantitative assay that does not account for preformed neutralizing complexes or valency and balance of circulating isotypes (2[IgG]-10 [IgM] sites) within samples. and the Abbott Panbio™ COVID-19 IgG/IgM rapid test device diagnostic assays [Choudhry et al. 2021; Gaeta et al. 2023], which gives a binary (+/-) indication of infection. A rabbit polyclonal anti-SARS-CoV-2 RBD was purchased (Stratech, Cat No 40592-T62-SIB).

### SARS-CoV-2 GloBodies

The polypeptide sequences of the published reference “Wuhan” strain [Zhou et al. 2020] were used to generate the SARS-CoV-2 GloBodies. (Figure 1A-1C). The DNA encoding these were synthesized and assembled into requested vectors by Twist Bioscience, California, USA. All sequences and plasmids are available upon request (ASK).

**Figure 1.**
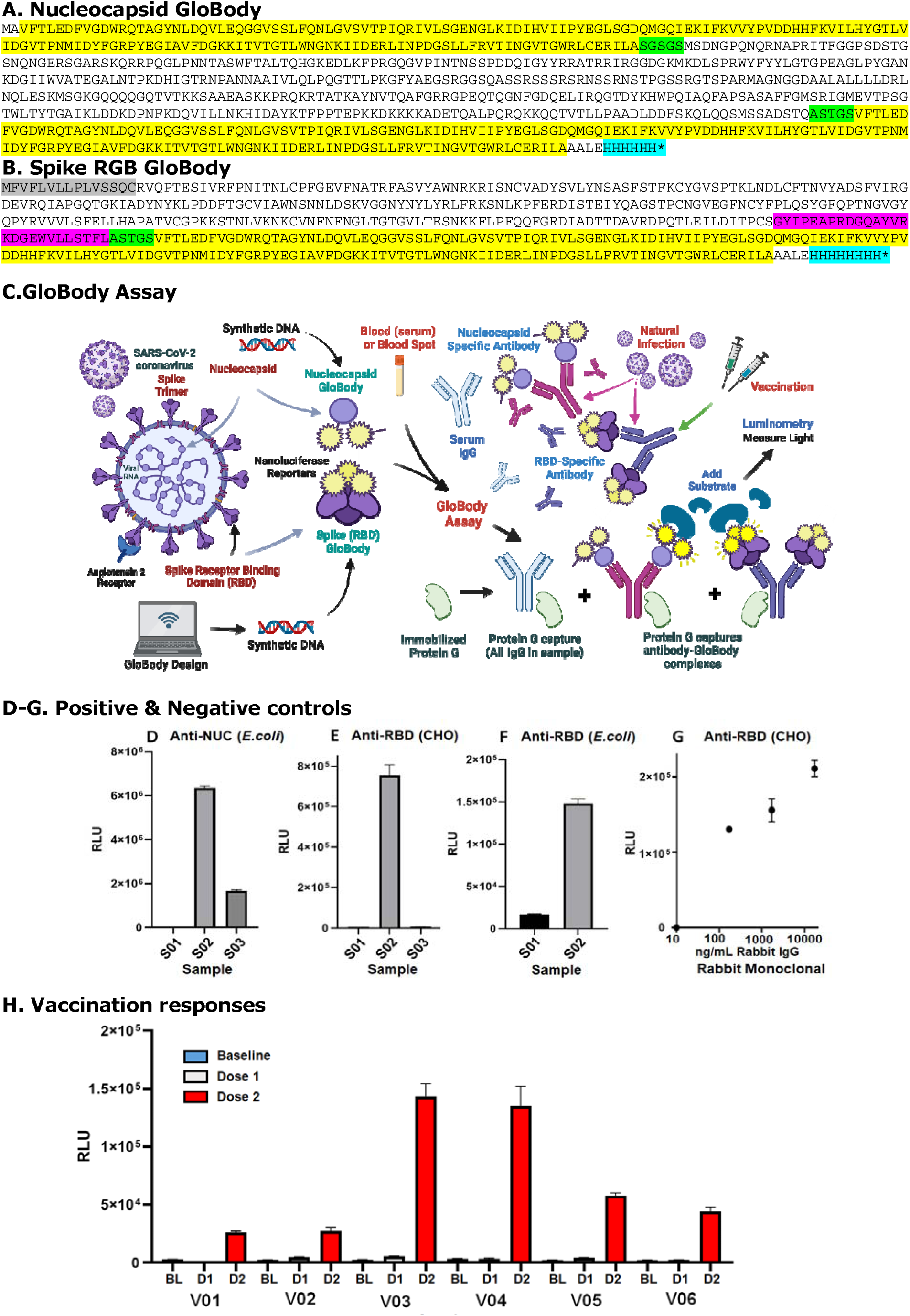
SARS-CoV-2 specific GloBodies and their Assay. The translated amino acid sequence of the (A) Nucleocapsid GloBody and (B) Spike RBD GloBody demonstrating the leader sequence (grey), linkers (Green), the T4 trimerization domain (magenta), nanoluciferase (yellow) and the histidine tags (cyan). These were used in (C) GloBody assays (Created in Biorender.com. https://BioRender.com/5h2brc1) to (D-F) detect nucleocapsid or RBD-specific antibodies in human blood spots. The RBD GloBodies were made in (D, F) bacterial (*E.coli*) cells and (E, G, H) mammalian (Chinese Hamster Ovary) cells. GloBody: antibody complexes were captured using protein G agarose and quantitated as relative light units (RLU) following the addition of furimazine substrate. Sample S001 was COVID-19 negative, S002 was hospitalised with COVID-19 and confirmed positive via PCR testing during the Wuhan strain infection wave (March 2020-August 2020). Sample P003 exhibited COVID-19- related symptoms but was not tested for PCR. (G) A rabbit polyclonal anti-SARS-CoV-2 RBD antibody was spiked into human sera and detected. (H) Sample V01-VO6 were taken at baseline and 4 weeks after vaccination with SAR-CoV-2 RNA (BNT162b2) and tested with the CHO-RBD GloBody made in mammalian cells. The results represent the mean and standard deviation of triplicate samples

### Nucleocapsid GloBody

The plasmid encoding SARS-CoV-2 nucleocapsid flanked with N and C terminal nanoluciferase reporters was designed for cytoplasmic expression in *Escherichia coli.* (Figure 1A). The recombinant protein was made in the cytoplasm of the *E.coli* BL21(DE3) strain following standard protocols [Studier 2005]. Aliquots of ∼1 x 10^8^ lux, light generating units, in 15 mM Tris pH8.0, 10% sucrose w/v and 2.5 mM MgCl_2_, were prepared and stored at -80°C until required.

### Spike RBD GloBodies

The Spike RBD GloBody construct was designed to be secreted into the culture media and codon optimised for Chinese Hamster Ovary (CHO) cell expression [Xu et al. 2020]. The RBD was linked with a T4 phage trimerization motif and a nanoluciferase reporter followed by a C-terminal octa-histidine tag (Figure 1B, 1C). This was assembled into pcDNA5/FRT for transfection Flp-In™-CHO (Invitrogen) cells following the manufacturer’s instructions. The medium was changed every 3-4 days until cell foci developed. These were detached and cloned by limiting dilution. The supernatants were tested for nanoluciferase activity. Cells were expanded into flasks the spent media was harvested/stored and GloBody was isolated using Ni^2+^NTA column affinity purification (Qiagen) Aliquots of ∼1 x 10^8^ lux, light generating units, in 15 mM Tris pH8.0, 10% sucrose w/v and 2.5 mM MgCl_2_, were prepared and stored at -80°C until required Subsequently, the RBD-GloBody was designed for cytoplasmic expression and assembled in the pET21b+ vector. This was transformed in *E.coli* NEB SHuffle® T7 Express and the recombinant protein affinity was purified as described before. Aliquots of ∼1 x 10^8^ lux, light generating units, in 15 mM Tris pH8.0, 10% sucrose w/v and 2.5 mM MgCl_2_, were prepared and stored at -80°C until required.

### GloBody Assay

Test samples were performed blinded and in triplicate. Four mm discs were punched (Rayher Hole Punch) from a single dried blood spot into 0.2 mL phosphate-buffered saline (PBS) containing 1% Triton X-100 and left overnight at ambient temperature. The sample was stored at -20°C, until required. For the eluted blood spot and serum samples, 20µL and 4 µL were used respectively in the GloBody assays (assuming a range of ∼10-15 µg/µL max 60 µg IgG). To dissociate antibody:antigen complexes 50 µL 0.1M glycine pH 2.7 was added. To ∼1 x 10^8^ lux, (a vast excess), GloBody activity units, 6 µL of 0.1M Tris pH 9.0 added, the volume adjusted to 0.6 mL with PBS with 0.05% Tween 20 (PBST) and added to the acidified sample (Patton et al. 2005). To the neutralised mix a 50% slurry of Protein G agarose equilibrated in PBS (30 µL, capacity ∼150 µg IgG) was added and placed on a rotating wheel for 60 min at ambient temperature. The mixture was transferred to 0.8 mL Micro-Spin Columns (Pierce/Thermofisher, Bishops Stortford, UK) and the excess reagent was removed by centrifugation at 200g for 1 min and the residual Protein G/IgG/GloBody complexes washed with PBST 750 µL (5X) with centrifugation at 200 x g between each wash and a final wash with PBS. To elute the retained GloBody 100 µL 0.1M Glycine pH2.7 was added to the agarose resin at ambient temperature and centrifuged at 16,200 x g for 1 min into a collection tube containing 12 µL 0.1M Tris pH 9.0. Neutralized eluate (30 µL) was added to 0.1 mL NanoGlo® substrate (Promega) (20 µL furimazine in 1 mL PBS pH 7.2 with 0.1% bovine serum albumin) in triplicate and after 10 min the luminescence was determined on a CLARIOstar plus plate reader, using the pre-set nanoluciferase setting [Saxena et al., 2020]. In the relative absence of positive standards, a qualitative readout, with reference to the limit of blank (LoB) may be used (Armbuster & Pry, 2008). In this study the value was defined by LoB = mean luminescence blank + 2.58(standard deviation blank). Values greater than this would suggest the presence of anti-SAR-CoV-2 antibodies with a confidence of 99%.

### Statistical Analysis

Descriptive summary statistics compared baseline characteristics between dental health-care workers, non-dental healthcare workers and other professions at the first study visit. Longitudinal analyses of nucleocapsid and Spike RBD titres were modelled using linear mixed models following log_10_ transformation of titres. Age, sex, occupation type, and performance of exposure-prone procedures were included as fixed effects. (Visit number/participant Identification) was included as a random effect to account for dependence of repeated measures for the same patient and the individual time course of titres for each patient. Analysis of symptom occurrence was modelled using generalised linear mixed models with a logit function. Age, sex, occupation type, and performance of exposure-prone procedures were included as fixed effects; participant identification was modelled as a random effect.

## RESULTS

### GloBody Production

Following the development of the COVID-19 pandemic, SARS-CoV-2 GloBodies were designed and synthesised (Figure 1A, 1B), and upon receipt of the GloBody plasmid, small-scale bacterial protein production was undertaken, and the anti-nucleocapsid assay was developed using samples eluted from dried blood spots (Figure 1C-1H). Using general basic laboratory equipment, sufficient reagent for ∼1,000,000 tests was generated from 1L cultures within about a week. The assays worked for serum samples (Figure 2). Some negative, pre-COVID samples sometimes gave signals, as has been commonly noted due to the cross-reactivity of nucleocapsid antibodies with pre-SARS-CoV-2 coronaviruses [Ng et al. 2020; Okba et al. 2020; Ma et al. 2021]. The anti-SARS-CoV-2 nucleocapsid-GloBody assay was used to test pre-vaccination sera assessed using subsequently developed diagnostic platforms [Choudhary et al. 2021]. The GloBody test compared well with the diagnostic Roche Elecsys® anti-SARS-CoV-2 Pan Ig assay (Figure 2A-D) and the Abbott Panbio™ COVID-19 IgG/IgM tests (Figure 2E-G). However, through the use of agarose-immobilized anti-human IgG (Figure 2B), IgM (Figure 2C) and IgA (Figure 2D) as a capture reagent, it allowed semi-quantification and greater granularity of the antibody response and could detect IgM and IgA dominated responses (Figure 2). Overall, the assay overall correlated well with the PCR test results (Figure 2E).

**Figure 2.**
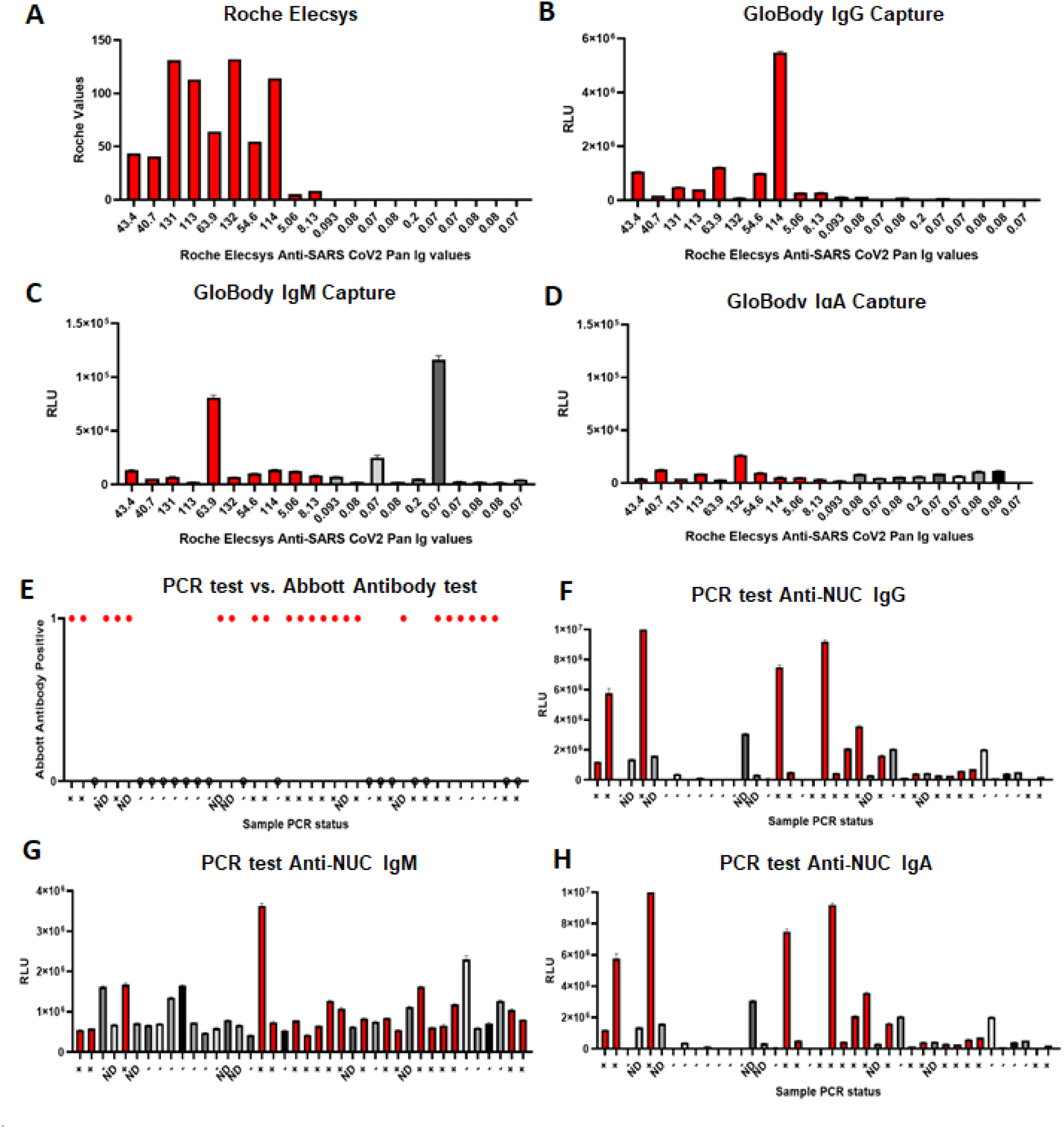
Nucleocapsid Globody compares well to diagnostic nucleocapsid-specific assays. Blinded serum samples were from those assayed on the diagnostic (A-D) Roche Elecsys® anti- SARS-CoV-2 Pan Ig assay or the (E-H) Abbott Panbio™ COVID-19 IgG/IgM Rapid Test anti- nucleocapsid lateral flow assay giving a binary positive or negative signal. Some had been confirmed infected using SAS CoV-2. These were designated as (A-D) positive (red) or (negative) based on Elecsys® assay or (E-H) antibody positive (Blue) or negative (grey) or PCR + antibody negative (green) based on the Panbio^TM^ assay. These were assayed with nucleocapsid GloBodies using agarose-immobilized anti-human (B, F) IgG (C, G) IgM or (D, H) IgA assays. The results represent the mean ± standard deviation of triplicate samples. N.D. No determined

The RBD GloBody reagent from mammalian cell culture could detect natural infection (Figure 1D), and as shown later, it detected vaccination responses (Figure 1H). The vaccine response was marginally but significantly (P=0.015 n=6) elevated 4 weeks after the first dose but became more marked (P=0.009. n=6) four weeks after the booster dose was administered up to 12 weeks later (Figure 1H). This was used to remotely monitor vaccination responses in people with multiple sclerosis using blood spots [Tallantyre et al. 2022]. Likewise, it could be used to monitor responses in animals since IgG from many species bind to protein G, and as an exemplar, responses were detected against a rabbit anti-SARS-CoV-2 antibody (Figure 1G) As proof of principle, the RBD GloBody could also be generated in bacterial cell cultures (Figure 1F).

### GloBody SARS-CoV-2 nucleocapsid and Spike-specific antibody responses in dental and medical HCW and scientists

Serial serum samples were collected from dental (n=82. Total 492 visits) and medical (n=72. Total of 432 visits) HCW and research laboratory-based scientists (n=62. Total 372 visits) (Table 1). Aerosol-generating dental procedures were suspended during the first national lockdown, and staff seconded to help the Medical HCW during the initial wave of ancestral “Wuhan-strain” SAR-CoV-2 and subsequent waves of COVID-19 affecting London (Figure 2A). Medical HCW performed exposure-risk procedures in 56.9% (n = 41/72) people before the first visit, but in 88.1% n= 317/360 cases before any of the visits where exposure risk was known. The majority of medical HCWs (95.8%. n=69/72) had experienced COVID-19- related symptoms at the time of the initiation of the study, and consistent with this, HCWs exhibited a consistent and higher nucleocapsid titres (Figure 3B). In contrast, it seemed that there were more dental HCW infections during the alpha SARS-CoV-2 infection wave that prompted the second and third National Lockdowns in late 2020/early 2021 (Figure 3A, 3B). Dental HCWs were highly exposed to viral-exposure-prone procedures (98.8% n=81/82 or 86.3% n=354/410 before any visit) (Table 1). Scientists were largely non-patient-facing individuals and they recruited later to the study as many were subject to “Stay at Home” conditions during the first National UK Lockdown. Few (6.5% n=4/62 before the first visit or 1.3% n=4/310 people before any of the visits) undertook viral-exposure-prone procedures (Table 1). The increased nucleocapsid GloBody titres of scientists may reflect acquisition from infection that was probably population-based rather than occupational-infection (Table 1, Figure 2B). The antibody titres became elevated as the two-metre distancing rule and opening of Blizard Building were relaxed and scientists seemed to show more elevated nucleocapsid-specific titres as the delta SARS-CoV-2 infection wave appeared (Figure 3A, 3B).

**Figure 3.**
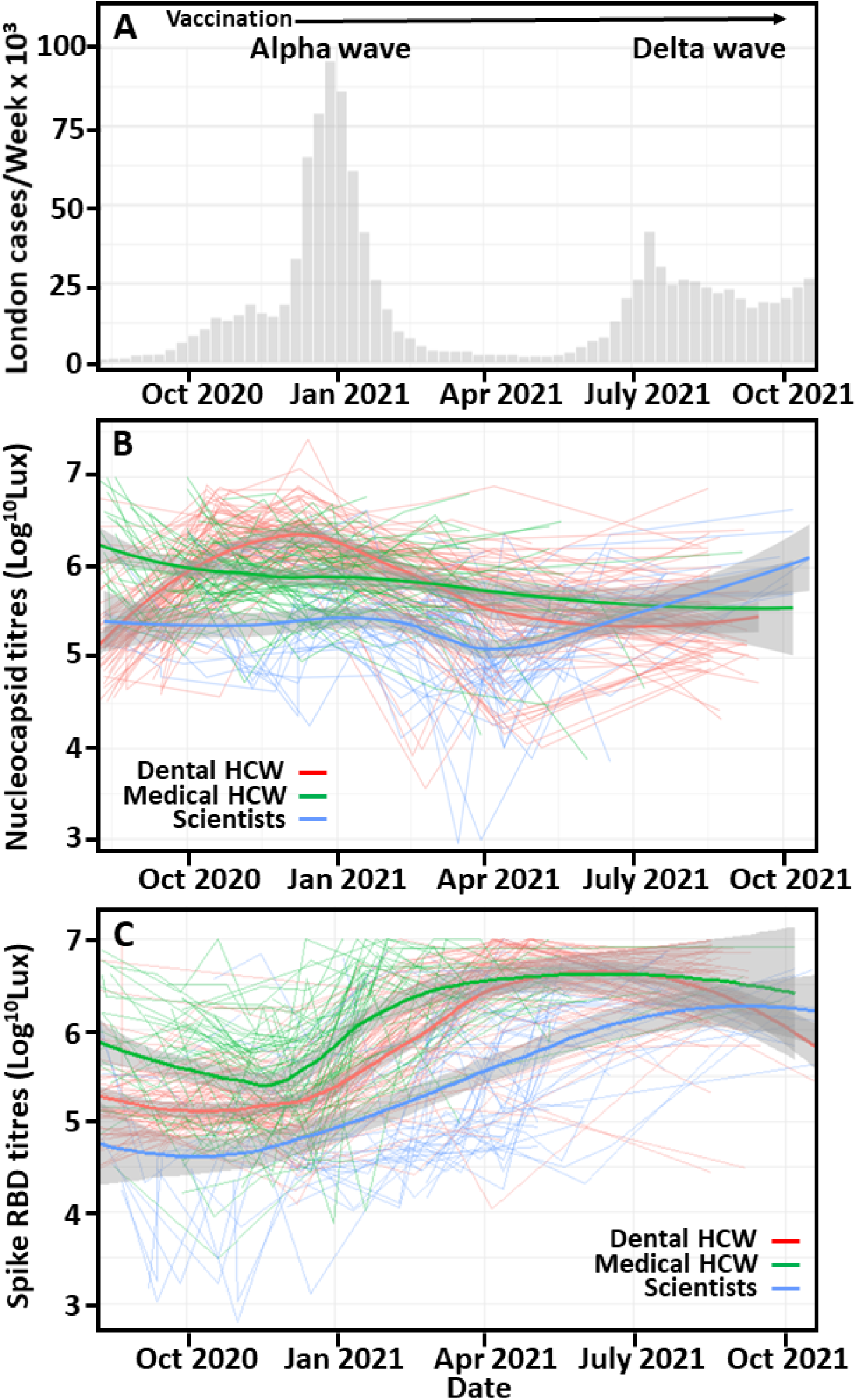
GloBody-detected trajectory of SARS-CoV-2-specific antibody responses in London-based dental healthcare workers, medical healthcare workers and scientists during the COVID- 19 pandemic. (A) The number of cases of COVID-19 per week in London during the period of sample collection were obtained from public weekly United Kingdom Government figures. (B, C) Serum samples were taken at monthly intervals from Royal London Hospital-based dental healthcare workers (HCW. Red), medical HCW (Green line) and scientists (Blue line) during the COVID-19 pandemic. These were assayed for: (B) Anti-nucleocapsid and (C) Anti-RBD IgG antibody responses using (B) nucleocapsid or (C) Spike RBD GloBody assays. GloBody:anti-viral antibody complexes were captured using protein G and quantitated following the addition of nanoluciferase substrate. The results represent detected light emitted (Lux units) from triplicate samples and show individual (thin lines), the mean (wide bold line) and standard deviation (grey).

**Table 1:** Characteristics of Dental Healthcare Workers, Medical Healthcare Workers and Scientists.

| Trait | Job Description |  |  | P value |
| --- | --- | --- | --- | --- |
|  | Dental | Medical | Scientist |  |
| **n** | 82 | 72 | 62 |  |
| Age (mean (SD)) | 38.5 (10.1) | 35.6 (10.0) | 35.2 (11.7) | 0.128 |
| Sex Male (%) | 22 (26.8) | 28 (38.9) | 17 (28.3) | 0.230 |
| Race/Ethnicity* (%) |  |  |  | <0.001 |
| Asian | 26 (31.7) | 10 (13.9) | 4 (6.5) |  |
| Black | 4 (4.9) | 6 (8.3) | 0 (0.0) |  |
| Mixed | 5 (6.1) | 0 (0.0) | 1 (1.6) |  |
| Other | 6 (7.3) | 5 (6.9) | 4 (6.5) |  |
| White | 41 (50.0) | 51 (70.8) | 53 (85.5) |  |
| Exposure prone procedures (%) | 81 (98.8) | 41 (56.9) | 4 (6.5) | <0.001 |
| Symptoms prior to first visit? (%) | 38 (46.3) | 69 (95.8) | 37 (59.7) | <0.001 |
| Time of first study visit (days) | 12.1 (13.7) | 52.0 (39.3) | 139.2 (67.6) | <0.001 |
| Time of first vaccination (days) | 162.6 (55.7) | 101.6 (43.9) | 60.8 (74.3) | <0.001 |
| First vaccination type (%) |  |  |  | <0.001 |
| Comirnaty®, Pfizer-BioNTech | 67 (81.7) | 56 (77.8) | 28 (45.2) |  |
| Vaxzevria®, AstraZeneca | 6 (7.3) | 2 (2.8) | 11 (17.7) |  |
| Spikevax®, Moderna | 1 (1.2) | 0 (0.0) | 2 (3.2) |  |
| Trial | 0 (0.0) | 0 (0.0) | 4 (6.5) |  |
| None | 8 (9.8) | 14 (19.4) | 17 (27.4) |  |
| Time of second vaccination(days) | 236.2 (50.9) | 176.0 (36.3) | 126.4 (72.6) | <0.001 |
| Second vaccination type (%) |  |  |  | <0.001 |
| Comirnaty®, Pfizer-BioNTech | 67 (81.7) | 36 (50.0) | 26 (41.9) |  |
| Vaxzevria®, AstraZeneca | 6 (7.3) | 0 (0.0) | 10 (16.1) |  |
| Spikevax®, Moderna | 1 (1.2) | 0 (0.0) | 2 (3.2) |  |
| Trial | 0 (0.0) | 0 (0.0) | 2 (3.2) |  |
| None | 8 (9.8) | 36 (50.0) | 22 (35.5) |  |

Demographics of recruited participants are reported as mean (standard deviation) or as a percentage of participants. The Time to first visit from initiation of study and time of vaccinations related to first study visit. The ethnicity was stratified to race/ethnicity according to the England and Wales census categories. Vaccines used during the early part of this study were all based around the Reference Wuhan strain of SARS-coV-2.

The Spike-RBD GloBody could detect natural infection and vaccination, as well as the decay in vaccination-induced antibody titres and the requirement for boosting to optimise vaccination responses (Figure 1H). In the HCW assessed here, the Spike-RBD serology was initially different and although elevated in dental and medical HCW compared to scientists (Figure 2C), the central driver for the rapid increase in titres was probably vaccination, which notably started to peak in early 2021. Although HCW/dental National Health Service staff and elderly people were prioritized for vaccination once available in December 2020. Most dental workers were vaccinated in January 2020 with a booster in March/April 2021. Scientific staff handling virus samples, processing and monitoring infection were given early access to available vaccines/vaccination capacity, including end-of-the-day, unused thawed RNA vaccine stocks, during the establishment of the East London vaccination centres. This more delayed access of scientists was evident in the RBD responses (Table 1. Figure 3A, 3C).

### Risk Factors for SARS-CoV-2 Antibody Responses

There was no apparent elevated risk of infection according to age or sex for nucleocapsid or Spike RBD GloBody responses (Figure 4A, 4B). However, there was a risk of elevated log_10_ nucleocapsid GloBody titres in non-dental, hospital-based HCW (0.37 95% CI 0.20-0.54 P<0.0001) and those people undertaking viral-exposure prone activities (0.21 95%CI 0.05- 0.38. P=0.012), indicative of higher infection levels (Figure 4A). Indeed, hospital-based HCWs, adjusted for vaccination status, had twice the odds of experiencing COVID-19 symptoms (2.01 95%CI 1.13-3.58. P<0.001) compared to dental HCWs. Likewise, those who performed virus exposure-prone procedures exhibited twice (1.98. 95% CI 1.18-3.34 P=0.01) the odds of symptoms. The levels of symptoms reported by dental workers were not different from research laboratory staff (1.46 95%CI 0.79-2.2.71. P=0.22). As expected, vaccination was associated with both reduced infection risk as shown by the reduced titre of nucleocapsid titres (-0.21 95%CI -0.34- -0.17. P<0.001. Figure 3A) and elevated Log_10_ RBD GloBody titres (1.18. 95%CI 1.09-1.26. P<0.001 Figure 1B). Compared to dental HCW, research laboratory staff exhibited reduced Log_10_ RBD GloBody titres (-0.3421 95%CI -0.54- -0.14. P<0.001. Figure 3B) and may reflect lower infection levels and delayed vaccination. There were no apparent differences in Log_10_ RBD GloBody titre in those undertaking exposure-prone procedures (0.03 95%CI -0.12-0.19. P=0.668) and those who do not, in contrast to that seen with nucleocapsid GloBodies. Furthermore, vaccinated people had half the odds of experiencing symptoms compared to unvaccinated individuals (0.51 95%CI 0.35-0.75. P<0.001), demonstrating the benefit from SARS-CoV-2 vaccination.

**Figure 4.**
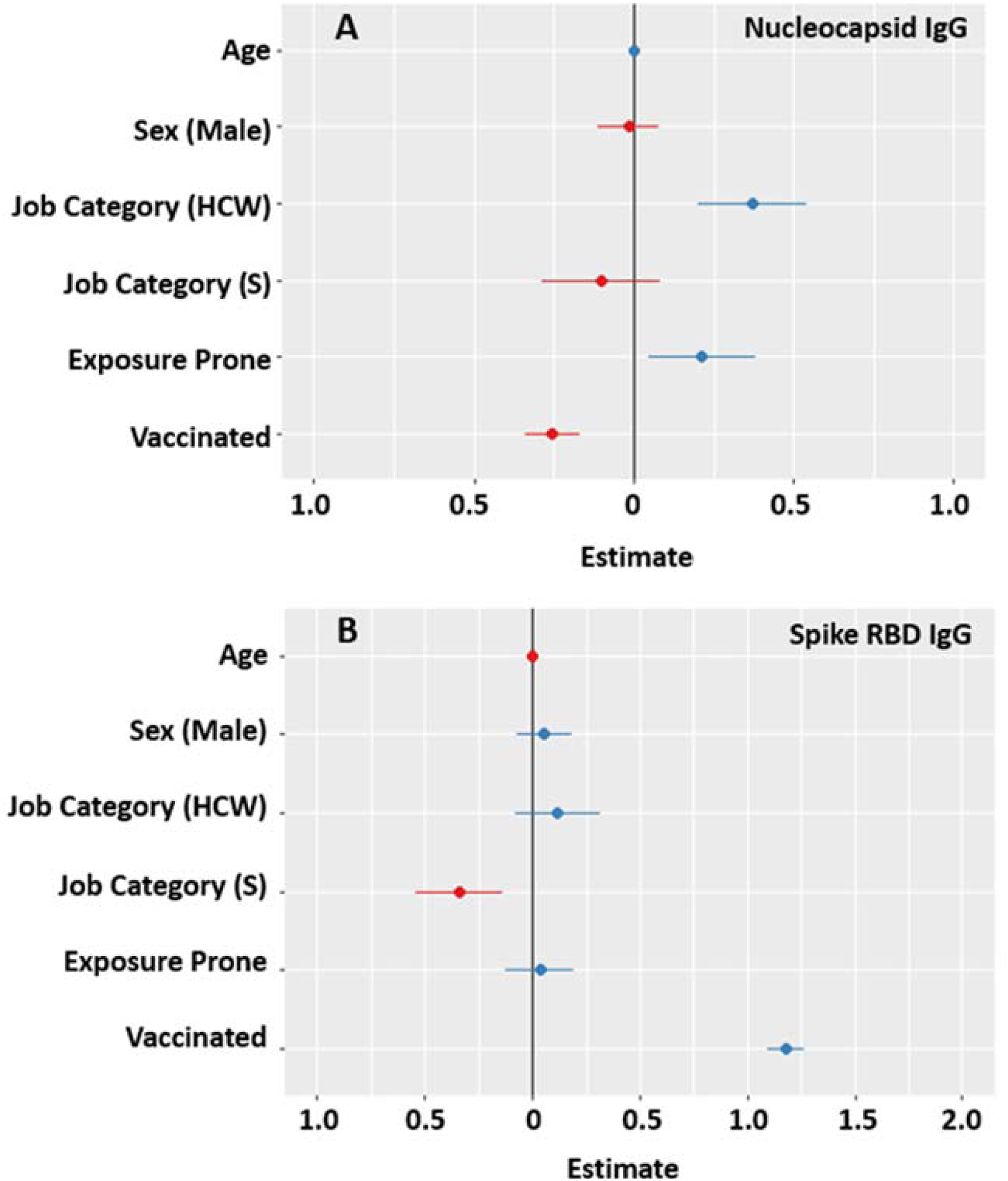
Factors associated with differences in anti-SARS-CoV-2 antibody titres. Serum samples were taken at two monthly intervals from hospital-based dental Workers (reference), hospital-based medical healthcare workers (HCW) and laboratory-based scientists (S) during the COVID-19 pandemic. These were assayed for: (A) Anti-nucleocapsid and (B) Anti-RBD IgG antibody responses using (A) nucleocapsid or (B) Spike RBD GloBody assays. Factors associated with the Log_10_ (A) Nucleocapsid- or (B) RBG-specific antibody GloBody titres were compared expressed by mean age of cohort, females, dentists, undertaking no exposure prone procedures prior to commencement of study and unvaccinated.

## DISCUSSION

East London, home to The Royal London Hospital, The Dental Institute, and the Queen Mary University, was at the heart of the COVID-19 outbreak in London. We examined the dynamics of exposure risk among staff at all three sites. Between 2020 and 2021, HCWs and those performing exposure-prone procedures faced the highest risk of COVID-19 exposure. In contrast, the research scientists examined here were relatively protected due to being subjected to the lockdowns implemented by the UK Government prior to the vaccine rollout. The COVID-19 public inquiry revealed that insufficient PPE stockpiles early in the pandemic, despite the importance of PPE, had been a recurring concern in previous preparedness planning exercises, including those for MERS-CoV, a related coronavirus [Hallett 2024]. The Department of Health & Social Care’s investigation into PPE supply during the pandemic, particularly in April and May 2020, found that a significant portion (at least 30%) of care workers, doctors, and nurses lacked sufficient PPE, even in high-risk settings [National Audit Office Report 2020]. The use of full PPE, notably-gloves, masks, gowns, and eye protection, was associated with a reduced risk of infection compared to partial or no PPE use [Scientific Advisory Group for Emergencies report 2021]

Indeed, other studies have shown that dentists and medical HCWs were at increased risk of infection [Magnusson et al. 2021; Van de Plaat et al. 2022], and the relative risks for COVID-19 among most patient-facing occupations were between 1.5 and 2.5 more than the general population [Plaat et al. 2022]. In the early part of the pandemic, the prevalence of SARS-CoV-2 in dental care workers was reported to be 11.8% (13,155/85,274; 95%CI, 7.5%-17%) and higher in dentists and dental hygienists than dental assistant personnel following meta-analysis of available literature [Schwarz et al. 2024]. Interestingly, the prevalence was lower 7.3% (95% CI, 5%-10%) in high-income countries compared to 20.8% (95% CI, 14%-29%; p<0.001) in low-middle income countries and adherence to the use of PPE was associated with lower SARS-CoV-2 prevalence rates [Srinidhi et al. 2023; Schwarz et al. 2024]. However, the data presented here suggests differing temporal exposure. Aerosol-generating clinical procedures were restricted in the dental profession at the start of the pandemic due to the need for adherence to stringent infection control protocols and fallow times following AGP procedures. This continued to affect clinical activity levels in the UK into 2022 when wide-scale vaccination took place. Thus, it is consistent with the data presented here. Medical HCWs were probably exposed to the reference ancestral strain when there was limited testing and restricted PPE prior to study commencement and exhibited higher titre nucleocapsid antibodies than dental HCWs and scientists at the start of the study. Although some dental HCWs were seconded to medical areas with higher COVID-exposure risks at the Royal London Hospital and elsewhere [Powell 2021], there was a notable rise in nucleocapsid titres in dental workers that corresponded with the period around the beginning of the second London lockdown as the population-infection of the alpha variant began to rise. This became problematic after people in London moved more freely. In contrast the small rise in laboratory researchers better coincided with the emergence of the delta variant, which exhibited some vaccine immune escape following vaccination before the second and third lockdowns [Bian et al. 2021]. The high titre of spike RBD antibody coincided with the introduction of SARS-CoV-2 vaccinations in HCW and the London population. However, with the easing of restrictions and the development of less lethal, vaccine-escape mutants such as omicron SARS-CoV-2 (B1.1.529 lineage) that became dominant in early 2022 [Willets et al. 2022], there were increased infections in the general population, which ceased to be actively and widely monitored in the UK in March 2023. It would be feasible to develop GloBodies to any SARS-CoV-2 variant rapidly.

The GloBody technology uses the generation of recombinant assay proteins. Due to advances and cost reduction in DNA synthesis, it was feasible to synthesise genes encoding whole molecules for tailored bacterial or mammalian expression. Bacterial cell expression allowed for rapid and easily scalable protein production that could quickly generate reagents for millions of assays. As the SARS-CoV-2 Spike protein is glycosylated [Xu et al. 2020], mammalian cell expression was originally selected for spike-related protein production. However, the reference strain RBD lacked important glycosylation, and therefore, the RGB GloBody could have been, and was subsequently also, produced by expression in *E.coli.* However, later strains, including alpha, delta and notably omicron SARS-CoV-2, exhibit extensive glycosylated RBD [Xu et al. 2020; Shajhan et al. 2023]. This demonstrates that the platform is flexible and can be optimised of either bacterial, strain-specific as needed, or mammalian cell expression. Although nucleocapsid and RBD-specific antibody expression was a product of SARS-CoV-2 infection, it was known early in the pandemic that there was some cross-reactive immunity from common cold-causing coronaviruses, influencing sensitivity of the available assays and many infections were asymptomatic [Ng et al. 2020; Okba et al. 2020; Ma et al. 2021]. Furthermore, time was required for the detection of viral-specific IgG and is spatially distinct from PCR-detected viral infection [Bunawan et al. 2021]. However, the nucleocapsid-related data in this study indicated that medical and dental workers exhibited evidence of infection/viral exposure prior to this occurring in scientists, suggesting an increased risk of occupational exposure.

This GloBody system has many advantages, notably the versatility and rapidity of generation, over a number of technologies developed to monitor SARS-CoV-2 infection in various university and commercial settings [Smithgall et al. 2020]. The SARS-COV-2 GloBody assays do not use live or infective viruses, and assay design and optimization can be performed under Biosafety category 1. Viruses within blood samples can be inactivated using detergents and/or heating [Darnell & Taylor 2006; Welch et al. 2020]. However, it soon became apparent that the UK National screening activity was funnelled into pre-formed, infection-related consortia with access to many negative and subsequent positive samples, often focussing on medical HCW [Martin et al. 2022]. Importantly, diagnostic companies selected national screening assays for use on their robotic diagnostic platforms within the health care setting. Thus, our focus was initially placed on remote monitoring of the patient population where testing was limited [Tallantyre et al. 2022]. Whilst diagnostic companies are more likely to develop high-volume tests, the GloBody system is versatile. It could be rapidly developed for a diagnostic platform or for rarer human or animal infections where commercial interest is currently lacking or not yet available.

In conclusion, our analysis of COVID-19 exposure across The Royal London Hospital, The Dental Institute, and the University in East London highlights the differentiated exposure risks among healthcare workers, dental professionals, and scientific staff during the pandemic. Findings demonstrated that frontline workers, particularly those in patient-facing roles, were at elevated risk of infection, compounded by PPE shortages, while lockdown measures initially shielded scientific staff. The study also underscores the efficacy of comprehensive PPE use in reducing infection risk and the impact of vaccination rollout on viral immunity markers. As demonstrated, the innovative GloBody technology offers a promising and versatile tool for detecting and monitoring SARS-CoV-2 and related antibodies, with potential application across various infectious diseases. Its rapid, scalable production capacity and adaptability for local and remote testing could provide an essential resource for global public health monitoring.

## Data Availability

Anonymized study data may be requested by qualified researchers from SG. Requests for unique reagents and methodology should be directed to ASK.

## Acknowledgements

The authors thank the students and staff for volunteering to participate in the study and thank the funders. We also thank people who supported our crowdfunding activities.

## Author contributions

Conceptualization: DB, SG, ASK, PLR. Data Curation: SG, UG, ASK, PK, TCM, PLR Ethics: DB, SG, PLR. Formal Analysis: DB, AH, ASK; Funding: GG, SG, ASK, PLR; Invention: ASK; Investigation-Assay: FA, GA, AG, SG, CH, ASK, AM, JS; Investigation- Resources: FA, MA1, MA2, DB, SG, GG, UG, ASK, DWH, CH, PK, EM, TCM, AP, PLR, RSR, KS; Methodology: ANA, DB, CC, RC, MJ, MRJ, ASK, ORM, TM, RSR; Project administration: DB, MA1, MA2, CH, SG, ASK, PLR; Visualization: DB, AH, ASK; Writing-Original Draft: DB, AH, SG, ASK, PLR; Writing-Review and Editing: All. All authors gave their final approval and agree to be accountable for all aspects of the work.

## Statements and Declarations

### Ethical Considerations

The study was conducted in accordance with applicable guidelines of Good Clinical Practice under UK Clinical Trial Regulations following ethical approval under Integrated Research Application System numbers IRAS 285071 and IRAS 284940 and Research Ethics Committee numbers 20/HRA/2743 and 20/NE/0176. Additional pre-vaccination, diagnostically-tested samples were obtained with ethical approval 20/HRA/2675, ISRCTN15634328.

### Consent to participate

Written informed consent was obtained from each participant.

### Consent for publication

No identifiable individual details are presented.

